# Association of neurotransmitter-based polygenic scores with antidepressant treatment outcomes in patients with major depressive disorder

**DOI:** 10.64898/2026.10.18.26365850

**Authors:** Nigussie T. Sharew, Scott R. Clark, Volker Arolt, Jürgen Brockmöller, Chia-Hui Chen, Katharina Domschke, Daniel K. Hall-Flavin, Ari Illi, Olli Kampman, Esa Leinonen, Ying-Jay Liou, Taisei Mushiroda, Julia C. Stingl, Amporn Benjaponpitak, Surakameth Mahasirimongkol, Liewei Wang, Masaki Kato, Yu-Li Liu, William V. Bobo, Shih-Jen Tsai, Michiaki Kubo, Richard M. Weinshilboum, David A. Mrazek, Joanna M. Biernacka, Bernhard T. Baune, Klaus Oliver Schubert, Azmeraw T. Amare

**Author notes:** **Correspondence to:** Azmeraw T. Amare, Adelaide Medical School, Adelaide University, Adelaide, SA 5000, Australia.

## Abstract

Biologically informed polygenic scores (PGSs) may help interpret genetic associations with treatment outcomes. Here, we computed neurotransmitter-based PGSs (*NeuroPGSs*) for patients with major depressive disorder (MDD) and assessed their association with SSRI treatment outcomes. By targeting genetic variants involved in key neurotransmission pathways, we developed PGSs for serotonergic (5228), gamma-aminobutyric acid (GABA, 4103), dopaminergic (7061), and glutamatergic (5690) variants. We leveraged data from two cohorts of patients who received SSRIs for at least 4 weeks: the International SSRI Pharmacogenomics Consortium (ISPC) and the Pharmacogenomics Research Network Antidepressants Medication Pharmacogenomics Study (PGRN-AMPS). Treatment response was defined as ≥50% reduction from baseline depression symptoms, measured using 17-item Hamilton Rating Scale for Depression (HRSD-17) in ISPC and 16-item Quick Inventory of Depressive Symptomatology (QIDS-C16) in PGRN-AMPS. Remission was defined as HRSD-17 ≤7 (ISPC) or QIDS-C16≤5 (PGRN-AMPS). The association between each *NeuroPGS* and antidepressant response or remission was evaluated using logistic regression models, adjusting for age, sex and five genetic principal components, with statistical significance determined at Benjamini–Hochberg-adjusted *p*<0.05. The GABAergic-based PGS was associated with SSRI response (aOR per SD = 1.45, 95%CI: 1.14-1.85). Patients with the highest GABAergic PGS (10^th^ decile) had 2.59-fold higher odds of response (95%CI: 1.46-4.60) compared to those with the lowest decile (1^st^ decile). These exploratory findings support further evaluation of *NeuroPGSs*; independent replication and assessment of incremental predictive value are needed before clinical application.

## 1. Introduction

Major depressive disorder (MDD) is among the leading causes of disability worldwide, affecting an estimated 300 million people in 2019 [1, 2]. Selective Serotonin Reuptake Inhibitors (SSRIs) are a major treatment component for patients with MDD [3, 4]; however, the number of patients for whom SSRI treatment is effective is limited. More than half of patients are treatment non-responders [5], and only about 30% of MDD patients show complete remission following the first drug prescribed [6, 7]. Non-responding patients often undergo several consecutive treatment steps, such as dose increase, switching to another antidepressant, or augmentation with different pharmacological agents [8, 9]. Genetic factors account partly to this variability, with one study estimating that common genetic variants explained 42% of antidepressant response variability (SE = 18 percentage points) [10], likely through complex and polygenic mechanisms [10, 11]. Pharmacogenetic studies have also identified associations between variation in the drug-metabolizing enzymes CYP2D6 and CYP2C19 and antidepressant plasma levels [12-14]. Despite these advances, limited tools are available to predict antidepressant response successfully, and no established predictive biomarkers or genetic signatures exist.

To date, genome-wide association studies (GWAS) aiming at identifying genetic markers for antidepressant treatment response have been largely unsuccessful [10, 11]. Given the complex nature of treatment outcome phenotypes, polygenic scores (PGSs) have been proposed as an alternative way to capture the cumulative effect of whole-genome genetic variants [15]. Thus, PGSs derived from MDD [16], schizophrenia (SCZ) [17], openness and neuroticism personality traits [18], SSRI responsiveness [11, 19], coronary artery disease [20] and obesity and cardioembolic stroke [21] have shown significant but modest associations with antidepressant response, explaining approximately 1% of interindividual response variability. However, a key limitation of genome-wide PGSs is their lack of biological resolution and no insight into how overall genetic load is apportioned among distinct biological pathways, which limits their interpretability and clinical translation.

To help overcome this limitation, biologically interpretable scores using a small list of genetic variants, such as pathway-specific PGSs that leverage variants involved in specific molecular processes, have been adopted [22-24]. These scores may enhance the predictive power of PGS models and have direct implications for pharmacogenomics or disease risk stratification [22]. For instance, our prior work showed that pathway-specific PGSs based on genes within acetylcholine, gamma-aminobutyric acid (GABA), glutamate, dopamine, calcium channel signalling, circadian rhythm, mitochondria, glycogen synthase kinase (GSK) and N-methyl-D-aspartate (NMDA), improved the prediction of long-term clinical response to the mood stabilizer lithium [23]. Similarly, PGSs developed from variants associated with nervous system development and neuron differentiation explained 6.9% of the variance in liability to psychosis, nearly doubling the 3.7% explained by trait-based genome-wide PGSs, in a sample of patients with DSM-IV diagnoses of schizophrenia or psychosis-related disorders [24].

Neurotransmitters such as serotonin, glutamate, gamma-aminobutyric acid (GABA), and dopamine play critical roles in the pathophysiology of depression [25-28]. Dysregulation of serotonin transmission has long been implicated; for example, reduced serotonin transporter binding has been reported in subsets of depressive patients [29, 30], while these findings remain inconsistent and effect sizes are modest [31]. Lower levels of glutamate, a major excitatory neurotransmitter that modulates postsynaptic excitability and neuroplasticity [32, 33], and reduced levels of GABA, a major inhibitory neurotransmitter responsible for overall control and fine-tuning of excitatory transmission, have been documented in depression [34-36]. Dopamine neurotransmission is implicated in modulating the motivation to seek pleasurable experiences, which is often reduced in depression [37, 38].

Given the role of neurotransmitters in depression pathophysiology, we sought to develop neurotransmitter-based PGSs (NeuroPGSs) to examine associations with antidepressant treatment outcomes. We also compared these associations with those of the conventional whole-genome PGSs for antidepressant responsiveness.

## 2. Methods and Materials

### 2.1. Study cohort characteristics

#### International SSRI Pharmacogenomics Consortium (ISPC)

The ISPC cohort was established to investigate the genetic factors contributing to variation in SSRI responsiveness [6]. A total of 865 participants who received SSRIs were recruited from seven sites across the USA, Germany, Thailand, Taiwan and Japan. Participants were started on citalopram 20 mg/day or escitalopram 10 mg/day, with the dose increased at the four-week visit to 40 mg/day for citalopram or 20 mg/day for escitalopram. All sites included in this analysis had received approval from their local ethical review board to participate in the consortium. The 17-item Hamilton Rating Scale for Depression (HRSD-17) was used to measure response to SSRIs treatment at baseline and follow-up visits at 4 weeks [39]. Treatment responders were defined as individuals with ≥50% reduction from the baseline in the HRSD-17 score, and patients with a HRSD-17 score ≤7 at four weeks were considered as remitters [6].

#### Pharmacogenomics Research Network Antidepressants Medication Pharmacogenomics Study (PGRN-AMPS)

The PGRN-AMPS cohort comprised 529 participants of European ancestry with non-psychotic MDD who were treated with fluoxetine, citalopram, escitalopram, or paroxetine (up to 40 mg/day). Clinical outcomes were assessed at four and eight weeks to examine treatment outcomes and genetic associations. Subjects, aged 18-84, were recruited from the Department of Psychiatry and Psychology at the Mayo Clinic in Rochester, Minnesota. Risks and benefits of the study were discussed with participants who gave written informed consent before entering the study, and the Institutional Review Board of the Mayo Clinic approved and monitored the treatment protocol. The baseline and follow-up at four 4 weeks assessments of depression severity were assessed using the 16-item Quick Inventory of Depressive Symptomatology (QIDS-C16) [40]. Treatment responders were those patients who showed at least a 50% reduction in QIDS-C16 score from baseline, while those with a QIDS-C16 score of 5 or less were categorized as remitters [41].

### 2.2. Genotyping, quality control, and imputation procedures

The genotype data for PGRN-AMPS were accessed via a controlled access system at the database of Genotypes and Phenotypes: dbGaP [42], while the ISPC data were obtained from the ISPC consortium [6]. Genotyping for ISPC cohort was conducted at RIKEN Center for Integrative Medical Science (Yokohama, Japan) using Illumina HumanOmniExpressExome BreadChips [6], whereas the PGRN-AMPS cohort was genotyped at the RIKEN Center for Genomic Medicine (Yokohama, Japan) using Illumina Human610-Quad BeadChips (San Diego, CA, USA) [43].

For both samples, we conducted quality control steps using PLINK version 2 [44] and samples with genotype call rates <95%, sex inconsistencies and genetically related individuals were excluded. Single-nucleotide polymorphisms (SNPs) that had a genotype call rate <95%, strand-ambiguous alleles (A/T and C/G), a minor allele frequency (MAF ≤1%) and SNPs with significant deviation from Hardy-Weinberg Equilibrium (HWE, *p* <10^-6^) were excluded in both cohorts. Genotype data passing QC criteria were imputed in the Michigan server separately for each study sample using the 1000 Human Genomes project reference panel. Detailed genotyping, quality control, and imputation procedures for both cohorts are provided in previous studies [6, 43].

### 2.3. Steps for developing neurotransmitter-based PGS (*NeuroPGS)*

#### Step one: Identification and mapping of genes and SNPs for each neurotransmitter

We selected four major neurotransmitter systems, i.e., serotonin [25, 28], dopamine [45], GABAergic [46, 47] and glutamatergic [34, 46] pathways, which are significantly implicated in depression aetiology and pharmacology. Genes were curated using Gene Set Enrichment Analysis (GSEA), HUGO Gene Nomenclature Committee (HGNC) and Kyoto Encyclopedia of Genes and Genomics (KEGG) gene databases by using neurotransmitters’ names as search terms (e.g., “dopamine”). After deduplication within each pathway, we retained 111 genes for serotonin, 76 for GABA, 87 for glutamate and 153 for dopamine. We used Multi-marker Analysis of GenoMic Annotation (MAGMA) software [48] to annotate SNPs to these genes in each neurotransmitter. SNPs were assigned to genes using the NCBI37 loc file with – *annotate window = 100, 20* (100kb upstream and 20kb downstream window). The final lists of annotated SNPs that were matched with our target dataset and included in our analysis were: serotonin (5228), GABA (4103), glutamate (5690) and dopamine (7061). The list of gene and SNP used in our analysis is provided in *Supplementary Tables 1-8*, and gene-set overlaps are shown in *Supplementary Figure 2*.

#### Insert supplementary materials here Step two: Compute NeuroPGS

We implemented a five-fold cross-validation procedure [49, 50], where *NeuroPGSs* were calculated for participants of one group at a time (target sample) using GWAS summary statistics from the remaining samples (discovery sample) in the combined cohort of PGRN-AMPS and ISPC cohorts. In each fold, 80% of the sample was used to generate GWAS summary statistics that were used as a discovery for PGS calculation in the 20% left-out target sample. The procedure was repeated five times by selecting a non-overlapping set of 20% left-out samples to calculate the PGS for the entire sample. Here, we generated five GWAS summary statistics for SSRI responsiveness and prepared five binary files for each split. Each GWAS was performed in PLINK with adjustment for age, sex, and the first five genetic principal components (PCs). A scree plot showing the number of PCs required to account for population structure in the combined cohort is presented in *Supplementary Figure 1*. In the original previous ISPC study, population structure was adequately captured by study site, with no evidence of residual population stratification within sites; therefore, no additional adjustment using ancestry PCs was required [6]. Polygenic scores were computed using the PRS-CS method, which incorporates continuous shrinkage priors on effect sizes and accounts for linkage disequilibrium (LD) among SNPs [51]. To assess the comparability of the cross-validation partitions, we provided the distribution of key demographic, ancestry, and treatment-outcome characteristics across the five folds *(Supplementary Table 9*). For comparison with the pathway-based scores, we developed a genome-wide PGS for antidepressant response using a five-fold cross-validation procedure and assessed its association with SSRI treatment outcomes. Further details on the development of *NeuroPGSs* are provided in the *supplementary material*.

#### Step three: Polygenic score association analysis

The association of each *NeuroPGS* with SSRIs treatment outcomes (Response/remission) was evaluated using a logistic regression model adjusted for age, sex and the first five PCs. A statistically significant association was determined at *p* ≤ 0.05 after multiple testing using the Benjamini-Hochberg Procedure (BH) [52]. The increment in Nagelkerke’s pseudo-R^2^ [53] was calculated as the difference between the model fit with specific PGSs and covariates and the model with only covariates. Stratified analysis was conducted by dividing the study subjects in both cohorts into PGS deciles, ranging from the lowest (1^st^ decile) to the highest (10^th^ decile) polygenic load for genes involved in each neurotransmitter. Then, the odds ratio of SSRIs treatment outcomes for patients with MDD who have a higher polygenic load for genes involved in neurotransmitters is compared to those in the lower PGS decile. We also performed ancestry-based sensitivity analyses to evaluate the associations of *NeuroPGS* and genome-wide PGS for antidepressant response with SSRI treatment outcome separately among participants of Asian and European ancestry. Figure 1 shows the development of PGSs for neurotransmitters and the detailed steps of the data analysis process.

**Figure 1:**
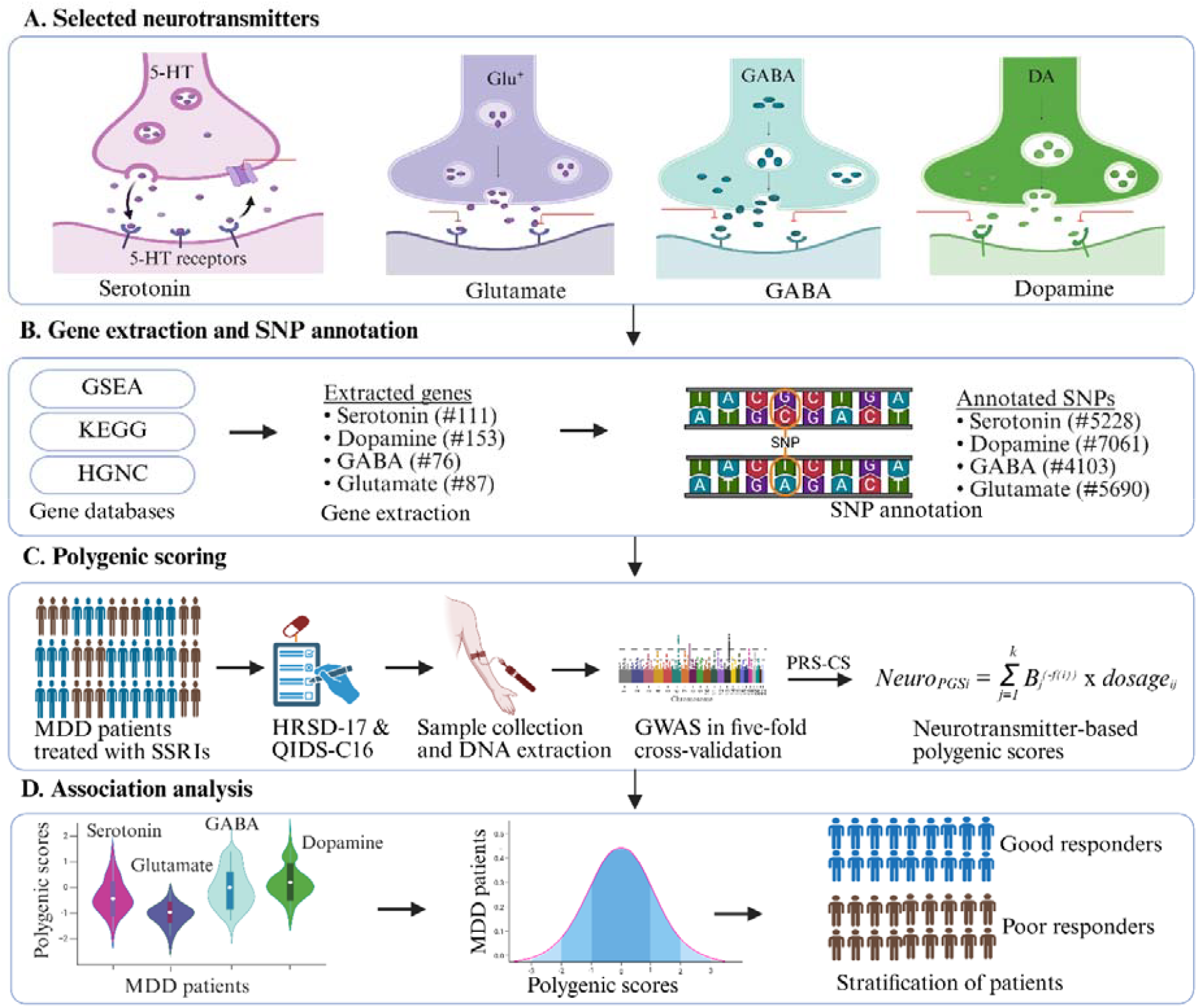
The development of PGSs for neurotransmitters and the detailed steps of the data analysis process. *Abbreviations:* 5-HT: Serotonin, Glu: Glutamate, GABA: Gamma-aminobutyric acid, DA: Dopamine, GSEA: Gene Set Enrichment Analysis, HGNC: HUGO Gene Nomenclature Committee, KEGG: Kyoto Encyclopedia of Genes and Genomes, SNP: Single-nucleotide polymorphism, MDD: Major depressive disorder, HRSD-17: 17-item Hamilton Rating Scale for Depression, QIDS: 16-item Quick Inventory of Depressive Symptomatology, PRS-CS: polygenic risk score with continuous shrinkage, NeuroPGS: Neurotransmitter-based polygenic score, PGS: Polygenic score

## 3. Results

### 3.1 Baseline characteristics of study participants

Our analysis included 865 patients with MDD from the ISPC and 529 MDD patients from the PGRN-AMPS pharmacogenomic cohorts who were treated with SSRIs. Nearly 65% of the participants were female, with a mean age (standard deviation) of 42.2 (14.5) years. Across both cohorts, 44.6% of study participants were categorized as responders, and 26.7% experienced remission.

### 3.2. Associations of neurotransmitter-based PGS and SSRIs treatment outcomes

We found a positive association between PGS computed from variants on genes involved in GABAergic neurotransmission (GABA_PGS_) and response to SSRIs in patients with MDD (aOR = 1.45, 95% CI: 1.14-1.85, *p* = 0.01; R^2^ = 1.25%; *Table 2*). Neither the association between the PGS for variants of genes involved in the serotoninergic pathway (Serotonin_PGS)_ and response (aOR = 1.12, 95% CI: 0.87-1.44, *p* = 0.37) nor that between the GABA_PGS_ and remission (aOR = 1.32, 95% CI: 1.00-1.74, *p* = 0.15) survived correction for multiple testing. In the decile-stratified analysis, MDD patients with the highest PGS decile (10^th^) in GABA_PGS_ had 2.59-fold higher odds of response to SSRIs (95%CI: 1.46-4.60) than those in the lowest decile (1^st^) *(Figure 2A)*.

**Figure 2:**
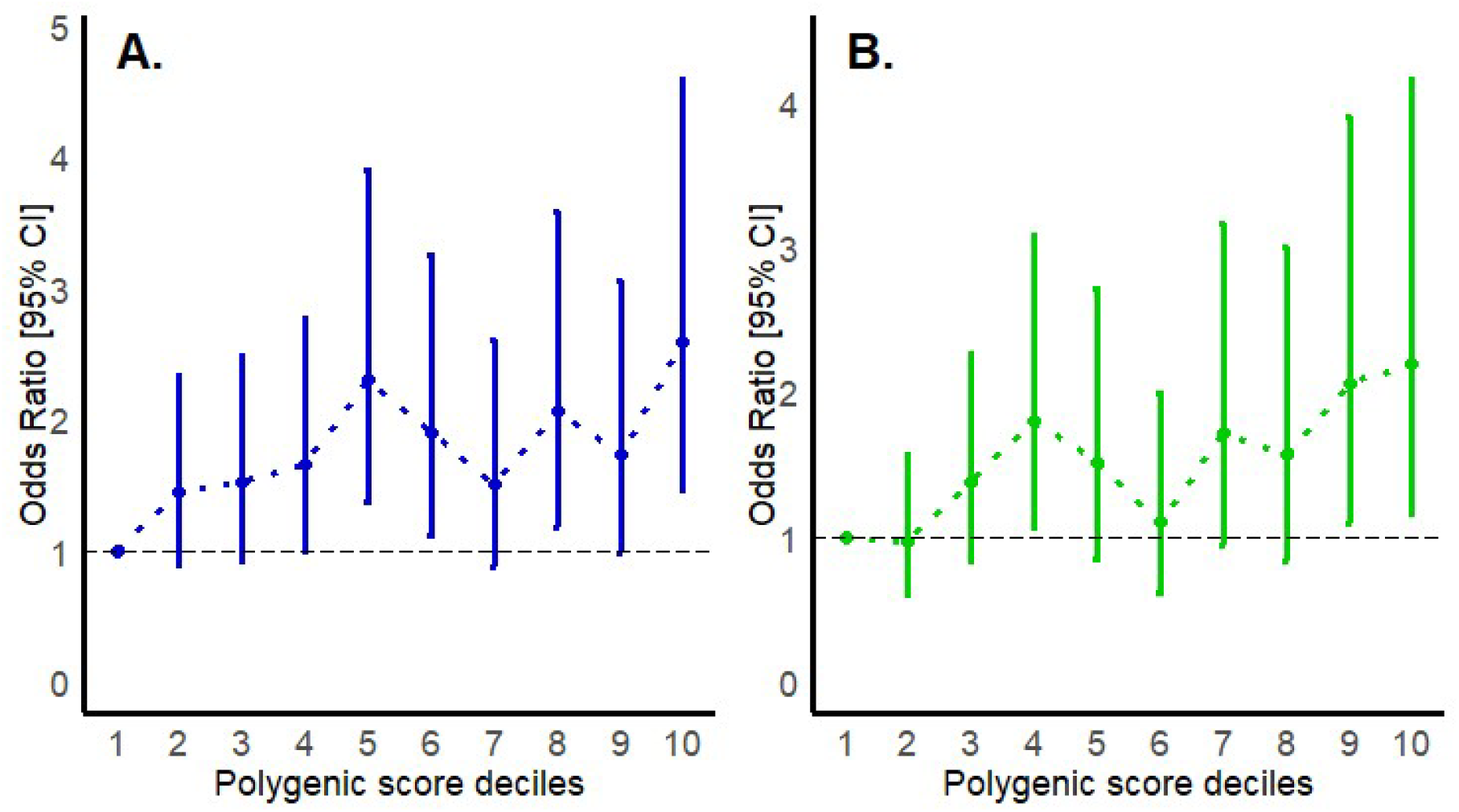
Trends in the odds ratios for good treatment response in patients with MDD, comparing those with a higher GABAergic (A) and genome-wide antidepressant responsiveness variants (B) PGS deciles (2^nd^ to 10^th^ ), with those patients with the lowest genetic score (decile 1^st^) (N□=□1394). *Legends:* The dot points and error bars represent the odds ratio and 95% confidence interval for the respective polygenic deciles. Confidence intervals that include an odds ratio of one are compatible with no association at the nominal 5% level. The analysis models were adjusted for age, sex, and the first five PCs *Abbreviation*: GABA = Gamma-aminobutyric acid

**Table 1.** Participant characteristics and outcomes after 4 weeks of SSRI treatment.

| Cohort | N | Sex, female,<br>N (%) | Age, Mean (sd) | Responders,<br>N (%) | Remitters,<br>N (%) |
| --- | --- | --- | --- | --- | --- |
| ISPC | 865 | 561 (64.9) | 43.7(14.7) | 416(48.1) | 226(26.1) |
| PGRN-AMPS | 529 | 335(63.3) | 39.6(13.7) | 206(38.9) | 128(24.2) |
| Total | 1394 | 896(64.3) | 42.2(14.5) | 622(44.6) | 354(26.7) |

*Abbreviations:* ISPC = International SSRI Pharmacogenomics Consortium; PGRN-AMPS = Pharmacogenomics Research Network Antidepressants Medication Pharmacogenomics Study; sd = standard deviation; N = Sample size

**Table 2.**
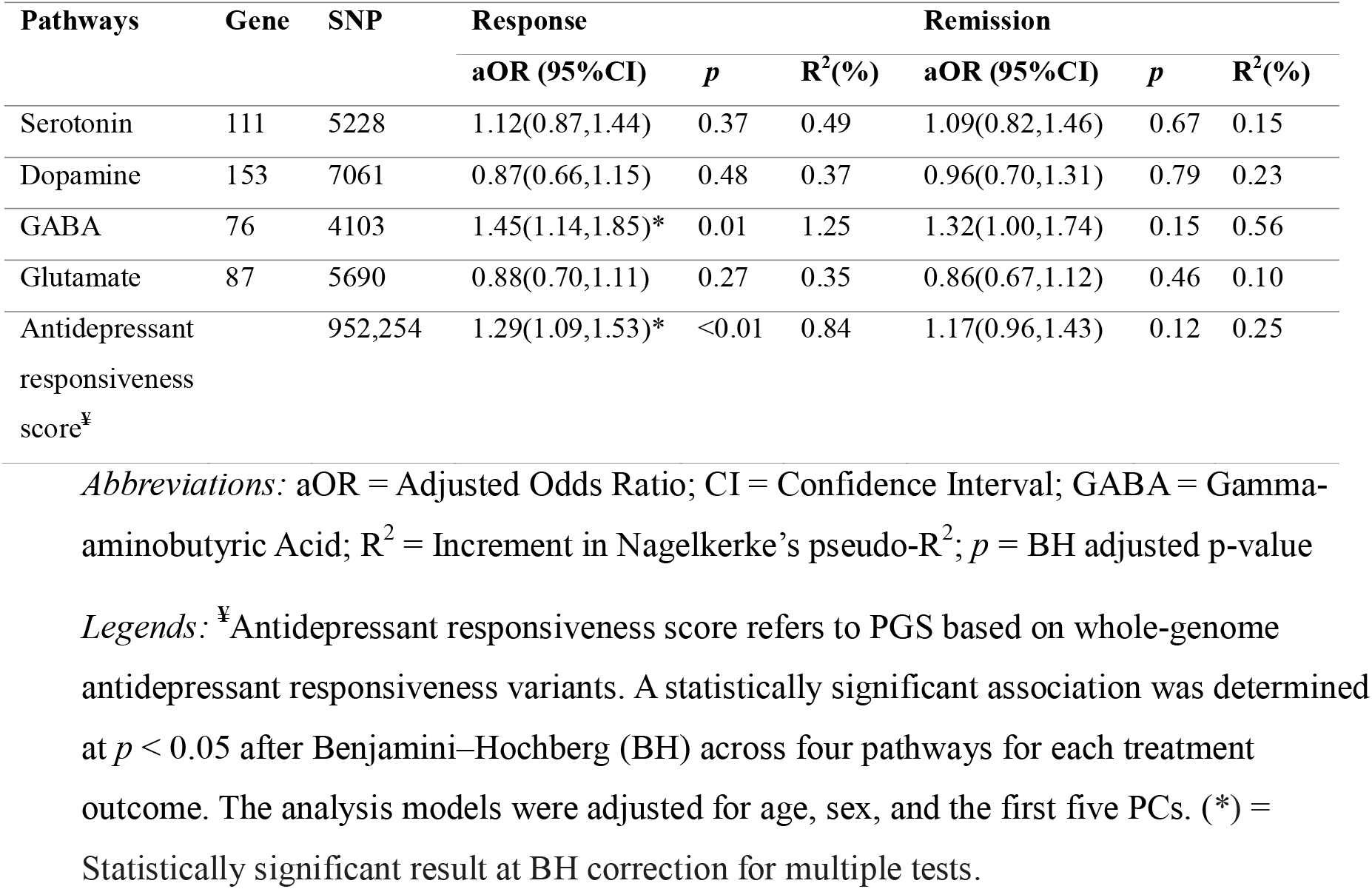
The association of neurotransmitter-based PGSs and SSRIs treatment outcomes.

We also found a positive association between the PGS for antidepressant responsiveness variants and response to SSRIs treatment (R^2^ = 0.84%, *p* < 0.01), but not with remission (*p* = 0.12). Patients with MDD who had a higher genetic load for antidepressant responsiveness variants had higher odds of being good responders to SSRIs treatment than those in the lower loads (aOR = 1.29, 95% CI: 1.09-1.53; *Table 2*). In stratified analysis, patients at the highest PGS decile (10^th^) for antidepressant responsiveness variants had 2.19-fold higher odds of response to SSRIs (95% CI: 1.15-4.16) compared to the lowest decile (1^st^) *(Figure 2B)*.

The association of the GABAergic pathway and overall antidepressant responsiveness variants polygenic scores with SSRIs response in stratified analysis across all deciles is provided in the *Figure 2 and Supplementary Table 10*.

### Sensitivity analysis

In ancestry-stratified analyses, the GABA_PGS_ was positively associated with SSRI treatment response among participants of Asian ancestry (aOR = 1.62, 95% CI 1.16-2.25; *p*=0.02). No neurotransmitter-specific PGS was significantly associated with remission in this group or with either outcome among participants of European ancestry. The genome-wide antidepressant response PGS was also positively associated with treatment response in the Asian subgroup (aOR = 1.28, 95% CI 1.05-1.58; *p*=0.02). Full association results for both ancestry groups are presented in *Supplementary Tables 11 and 12*.

## 4. Discussion

This study investigated the association of pathway-specific polygenic scores with SSRIs treatment outcomes in patients with MDD, with a particular focus on genetic variation within key neurotransmitter systems, including serotonergic, dopaminergic, GABAergic, and glutamatergic pathways. Our results indicated that a higher response-weighted GABAergic PGS was positively associated with SSRIs response (aOR = 1.45, 95% CI: 1.14-1.85). Individuals in the highest decile (10^th^ decile) of the GABAergic PGS had more than two-fold higher odds of a favourable response compared with those in the lowest decile (1^st^ decile).

The association between GABAergic PGSs and SSRI treatment response was in the same direction as findings derived from the conventional genome□ wide PGS for antidepressant responsiveness. For instance, in the current study, we observed that individuals with higher genetic load for genome-wide variants of antidepressant responsiveness had 1.3-fold higher odds of response to SSRI treatment than those with lower genetic load. Similarly, a study by Men et al. found that depressed patients treated with sertraline and olanzapine who had a higher genetic load for antidepressant responsiveness had nearly two-fold higher odds of reaching remission [54]. In terms of explained variance, GABAergic-based PGS was associated with an increment of approximately 1.3 percentage points in Nagelkerke’s pseudo-R^2^, which is comparable with genome-wide PGS derived from GWAS of MDD remission and percentage improvement in a larger antidepressant cohort (N=5218; R^2^ = 0.8%) [11]. These associations do not establish that the GABAergic PGS provides greater predictive value than the genome-wide score.

The observed positive association between a response-weighted GABAergic PGS and SSRI response is also consistent with a substantial body of prior evidence implicating GABA dysfunction in the pathophysiology of depression and in antidepressant treatment mechanisms. Numerous studies have demonstrated that GABA levels are reduced in patients with depression, both centrally in the brain [55-58] and peripherally in plasma [59], and that these levels tend to be higher in antidepressant responders compared with non□ responders [33]. Furthermore, antidepressant treatment has been shown to normalize GABA levels [60, 61], and alterations in GABAergic transmission represent a core component of the pathophysiological cascade underlying MDD that can be reversed by monoaminergic antidepressant action [47, 59]. At the neurobiological level, post□ mortem and imaging studies have reported reduced expression of GABA□ synthesizing enzymes [62, 63], a decreased density of GABAergic interneurons [64], and reduced expression of specific GABAergic type A α4 and *δ* receptor subunits [65] in the brains of individuals with MDD. Collectively, both clinical and preclinical evidence support the notion that deficits in GABAergic inhibition contribute to depressive symptoms and influence antidepressant responsiveness [66, 67].

Given the delayed onset of action and limited efficacy of conventional SSRIs, there has been increasing interest in developing novel and rapid□ acting antidepressants that directly target GABAergic mechanisms [46, 68]. Benzodiazepines, which modulate synaptic GABAergic signalling by binding to the α–γ subunit interface of the GABA receptor, have shown therapeutic utility in improving early depressive symptoms when used as adjuncts to antidepressants [69]. Similarly, neuroactive steroids have emerged as a class of agents with rapid antidepressant effects in some clinical settings through their potent modulation of GABAergic and glutamate signalling [70, 71]. Preclinical studies have shown that the naturally occurring neurosteroid allopregnanolone is a positive allosteric modulator of synaptic and extrasynaptic GABA_A_ receptors, enhancing both phasic and tonic inhibition through increased chloride influx, neuronal hyperpolarization, and reduced excitability [72, 73]. Consistently, reduced cerebrospinal fluid levels of allopregnanolone have been reported in depression and normalize following antidepressant treatment [74]. Building on this evidence, GABAergic neurosteroid-based agents such as, brexanolone, ganaxolone and zuranolone have been developed and shown rapid antidepressant efficacy in clinical trials of postpartum depression [75-77], supporting the translational relevance of GABA modulation in depression.

Similar to our findings in MDD, higher genetic loading for GABAergic pathway variants has also been positively associated with treatment response to lithium in patients with bipolar disorder [23]. Modulation of the GABAergic transmission has been implicated in the mechanism of action of both antidepressants and mood stabilizers [78], suggesting potentially shared biological mechanisms across mood disorders [79]. This overlap has important implications for the development of therapeutic strategies targeting the GABAergic system across diagnostic boundaries.

Several limitations of our study should be addressed in the future. *First*, the reported significant association between GABAergic-based PGS and the treatment outcome was identified using a relatively small sample size [15]; this finding has yet to be replicated in larger independent cohort. *Second*, although scores were derived using GWAS within a five-fold cross-validation to minimize circularity[22], association analyses were conducted within the same pooled sample, which also warrants external replication. *Third*, the extracted genes may not exhaustively represent the neurotransmitter systems implicated in depression pathophysiology, as gene annotation depends on the current gene databases curation and may not fully incorporate all genes involved in each neurotransmitter [80]. *Fourth*, the observed gene overlap across the four neurotransmitters indicates partial dependence among PGSs, warranting caution when interpreting associations and multiple-testing correction. *Fifth*, while individuals of Asian and European ancestry are included in our study, further inclusion of cohorts from diverse populations helps to ensure the findings apply to worldwide populations. *Sixth*, the use of a European-ancestry LD reference panel may not fully capture LD patterns across participants with Asian ancestry, potentially affecting PGS estimation. *Seventh*, treatment outcomes were assessed after only four weeks of SSRI treatment, which may not capture patients who require longer treatment periods to achieve response or remission. *Lastly*, because all participants received SSRIs, these associations cannot distinguish general prognosis from SSRI-specific treatment benefit.

## 5. Conclusion

A GABAergic pathway-based PGS was associated with SSRI response in an internally cross-validated analysis. Independent replication and further evaluation of pathway specificity and incremental predictive value are needed before clinical application.

## Supporting information

Supplementary material: Genes and SNPs list by pathways

## Data Availability

All data produced in the present study are available upon reasonable request to the authors

## Acknowledgment

The authors are grateful to the study subjects who participated in the studies, and we appreciate the contributions of research staffs who helped in the patient recruitment and data collection for the studies. The authors would also like to thank the National Institutes of Health (NIH), USA for making the PGRN-AMPS accessible to us. The complete clinical data for the ISPC is available at http://www.pharmgkb.org/downloads/. The PGRN-AMPS data were obtained through controlled access distributed from the NIH in the dbGaP (https://www.ncbi.nlm.nih.gov/gap). The analysis of this study was carried out using the high-performance computational (HPC) capabilities of Adelaide University’s Phoenix Supercomputer https://staff.adelaide.edu.au/research-and-innovation/access-research-infrastructure/hpc-and-digital-infrastructure/phoenix-hpc

## Author contributions

AT Amare conceived and designed the project and secured a fellowship to lead the study. NT Sharew developed the research proposal, conducted the statistical analysis, interpreted the findings, and drafted the manuscript. AT Amare, SR Clark and KO Schubert provided supervision and critically reviewed the data analysis steps and manuscript draft. All authors contributed to cohort recruitment and writing and editing of the manuscript.

