## Supplementary material for "Association of neurotransmitter-based polygenic scores with antidepressant treatment outcomes in patients with major depressive disorder"

### Procedures for developing neurotransmitter-based polygenic scores

We partitioned the combined International SSRI Pharmacogenomics Consortium (ISPC) and the Pharmacogenomics Research Network Antidepressants Medication Pharmacogenomics Study (PGRN-AMPS) cohorts into five groups. Using this five-fold cross-validation approach, we generated 5 sets of GWAS summary statistics, each leaving one group out as the target and using the remaining groups as the discovery dataset for treatment response. Each GWAS was performed in PLINK version 2.0 with adjustment for age, sex, and the first five genetic principal components (PCs). The number of PCs was determined by examining the scree plot of eigenvalues in the principal component analysis; the first five PCs were selected to represent population structure (Supplementary Figure 1).

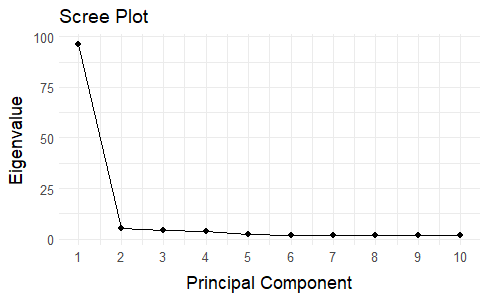

***Supplementary Figure 1:*** Scree plot for determining the number of genetic principal components (PCs) considered in our analysis

We used the polygenic risk score with continuous shrinkage priors (PRS-CS) method version 1.1.0, which has shown superior predictive performance over clumping and thresholding and other methods[1, 2]. PRS-CS employs a Bayesian regression framework, incorporating continuous shrinkage priors on effect sizes and accounting for linkage disequilibrium (LD) among SNPs[2]. Continuous shrinkage applies a proportionate amount of reduction to the coefficient estimates of SNPs based on their strength of association. This process yields posterior SNP effect estimates for downstream scoring while accounting for LD. To estimate SNP posterior effect sizes, genetic data from the UK Biobank European subsample was used as an external panel of reference for LD pattern and global shrinkage parameter *(--phi=1e-2*) with other default options, considering the highly polygenic nature of antidepressant treatment outcome and lower sample size[3, 4]. Polygenic scores for each neurotransmitter were then computed by summing up dosages of effect allele (ranging from 0 to 2) for SNPs (j to k) annotated within specific pathway, multiplied by their respective posterior SNP weight inferred from PRS-CS:

$$Neuro{PGS}_{i} = \sum_{j = 1}^{k} \hat{\beta}_{j}^{(-f(i))} \times{dosage}_{ij}$$

Here, i denoted the individual; j each SNP and k the number of scored variants in each neurotransmitter pathway. dosageᵢⱼ is the effect-allele dosage for individual i at SNP j and $\hat{\beta}_{j}^{(-f(i))}$ is the posterior SNP effect estimated excluding the fold containing individual i.

The procedure was repeated until every participant had an out-of-fold score for each neurotransmitter pathway. Prior to conducting association analyses, the scores for each neurotransmitter pathway were standardized within each fold to a mean of zero and a standard deviation of one. Table 1 below reports participant characterization across each fold. A major assumption in polygenic score computation is the absence of sample overlap between the discovery and target datasets, and this condition was checked in our study.

### Gene overlap across the four neurotransmitter pathways

The four neurotransmitter pathways showed both unique and shared genes. Most genes were pathway-specific, including 90 dopamine, 49 glutamate, 54 serotonin, and 49 GABA genes. The largest overlap was observed between dopamine and serotonin (48 genes), followed by glutamate and GABA (18 genes) and dopamine and glutamate (7 genes). Smaller overlaps were observed across other pathway combinations, indicating partial sharing of genes among the neurotransmitter systems *(Supplementary Figure 2).*

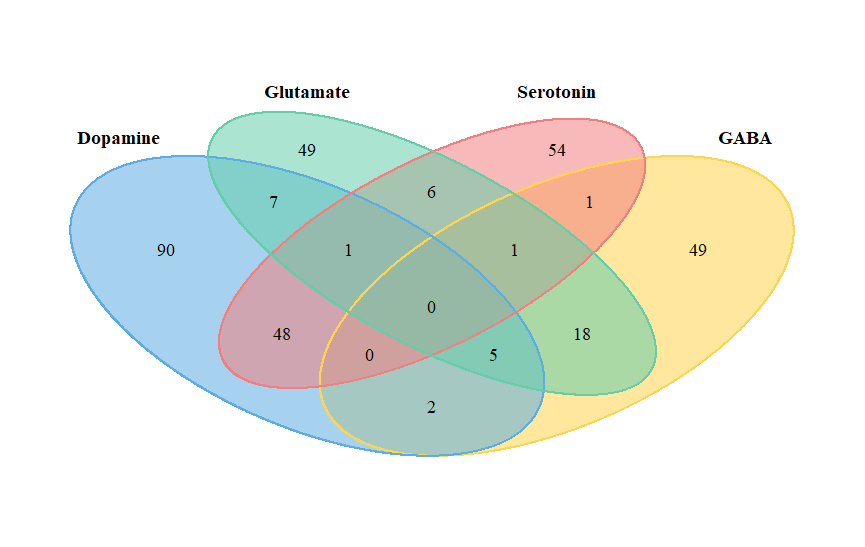

***Supplementary Figure 2:*** Gene overlap across the four neurotransmitter pathways

### Participant characteristics across each fold

***Supplementary Table 9***: Participant characteristics across each fold

| Folds | Total participants | Sex, female, N (%) | Age, Mean (sd) | Responders, N (%) | Remitters, N (%) | Asian ancestry N (%) |
| --- | --- | --- | --- | --- | --- | --- |
| 1 | 286 | 173(60.0) | 42.1(14.9) | 124(43.4) | 73(25.5) | 109(37.1) |
| 2 | 281 | 187(66.5) | 42.3(13,8) | 125(44.4) | 61(21.7) | 118(42.0) |
| 3 | 281 | 180(64.1) | 42.1(14.1) | 120(42.7) | 65(23.1) | 121(43.1) |
| 4 | 269 | 181(67.3) | 42.3(14.8) | 121(45.0) | 69(25.7) | 111(41.2) |
| 5 | 277 | 175(63.2) | 42.1(14.5) | 132(47.7) | 86(31.0) | 108(39.0) |
| Combined | 1394 | 896(64.3) | 42.2(14.5) | 622(44.6) | 354(26.7) | 567(40.7) |

### Stratified analysis of GABAergic-based and genome-wide PGSs with good SSRI response

**Supplementary Table 10:** The full association of the GABAergic pathway and genome-wide antidepressant responsiveness polygenic scores with good SSRIs response among patients with MDD in stratified analysis

| **Polygenic score decile** | **GABA_PGS_ (aOR; 95%CI)** | **Antidepressant responsiveness score (aOR; 95%CI)** |
| --- | --- | --- |
| 1^st^ | 1 | 1 |
| 2^nd^ | 1.44(0.89,2.34) | 0.97(0.60,1.56) |
| 3^rd^ | 1.52(0.92,2.49) | 1.38(0.84,2.62) |
| 4^th^ | 1.66(0.99,2.77) | 1.80(1.05,3.08)* |
| 5^th^ | 2.31(1.36,3.90)* | 1.51(0.84,2.71) |
| 6^th^ | 1.90(1.11,3.25)* | 1.10(0.61,1.99) |
| 7^th^ | 1.51(0.87,2.61) | 1.72(0.94,3.16) |
| 8^th^ | 2.06(1.18,3.59)* | 1.57(0.83,2.99) |
| 9^th^ | 1.73(0.98,3.05) | 2.06(1.09,3.90)* |
| 10^th^ | 2.59(1.46,4.60)* | 2.19(1.15–4.16)* |

*Legends:* *Legends:* **^¥^**Antidepressant responsiveness score refers to PGS based on whole-genome antidepressant responsiveness variants. A statistically significant association is determined at p < 0.05, adjusted for age, sex and the first five genetic PCs. (*) denotes statistical significance in the decile comparison of the highest genetic loads (10^th^ decile) with the lowest genetic loads (1^st^ decile).

*Abbreviations:* GABA: Gamma-aminobutyric acid

### Sensitivity association analysis of PGSs with SSRI treatment outcomes by ancestry

**Supplementary Table 11:** The association of neurotransmitter-based and genome-wide antidepressant response PGSs with SSRIs treatment outcomes among participants with Asian ancestry *(N = 567).*

| Polygenic score | Response | | | Remission | | |
| --- | --- | --- | --- | --- | --- | --- |
|  | **aOR (95%CI)** | ***p*** | **R^2^(%)** | **aOR (95%CI)** | ***p*** | **R^2^(%)** |
| Serotonin | 0.98(0.70,1.39) | 0.95 | 0.96 | 1.2(0.80,1.88) | 0.43 | 0.59 |
| Dopamine | 0.91(0.63,1.31) | 0.81 | 1.09 | 0.90(0.57,1.42) | 0.65 | 0.64 |
| GABA | 1.62(1.16,2.25)* | 0.02 | 1.78 | 1.48(0.98,2.22) | 0.25 | 1.33 |
| Glutamate | 0.87(0.61,1.25) | 0.81 | 1.10 | 0.76(0.49,1.18) | 0.43 | 0.70 |
| Antidepressant responsiveness score^¥^ | 1.28(1.05,1.58)* | 0.02 | 1.32 | 1.22(0.95,1.57) | 0.12 | 0.87 |

*Legends:* **^¥^**Antidepressant responsiveness score refers to PGS based on whole-genome antidepressant responsiveness variants. A statistically significant association was determined at *p* < 0.05 after Benjamini–Hochberg (BH) across four pathways for each treatment outcome. The analysis models were adjusted for age, sex, and the first five PCs. (*) = Statistically significant result at BH correction for multiple tests. *Abbreviations:* aOR = Adjusted Odds Ratio; CI = Confidence Interval; GABA = Gamma-aminobutyric Acid; R^2^ = Increment in Nagelkerke’s pseudo-R^2^; *p* = BH adjusted p-value

**Supplementary Table 12:** The association of NeuroPGS and genome-wide PGS antidepressant response with SSRIs treatment outcomes among participant with European ancestry *(N = 827).*

| Polygenic score | Response | | | Remission | | |
| --- | --- | --- | --- | --- | --- | --- |
|  | **aOR (95%CI)** | ***p*** | **R^2^(%)** | **aOR (95%CI)** | ***p*** | **R^2^(%)** |
| Serotonin | 1.31(0.91,1.88) | 0.42 | 0.22 | 1.02(0.68,1.52) | 0.67 | 0.03 |
| Dopamine | 0.85(0.53,1.36) | 0.49 | 0.01 | 1.02(0.61,1.71) | 0.37 | 0.01 |
| GABA | 1.27(0.87,1.86) | 0.42 | 0.68 | 1.23(0.81,1.86) | 0.33 | 0.11 |
| Glutamate | 0.85(0.62,1.17) | 0.42 | 0.01 | 0.89(0.63,1.26) | 0.51 | <0.01 |
| Antidepressant responsiveness score^¥^ | 1.33(0.96,1.85) | 0.90 | 0.51 | 1.11(0.78,1.59) | 0.12 | 0.06 |

*Legends:* **^¥^**Antidepressant responsiveness score refers to PGS based on whole-genome antidepressant responsiveness variants. A statistically significant association was determined at *p* < 0.05 after Benjamini–Hochberg (BH) across four pathways for each treatment outcome. The analysis models were adjusted for age, sex, and the first five PCs. (*) = Statistically significant result at BH correction for multiple tests. *Abbreviations:* aOR = Adjusted Odds Ratio; CI = Confidence Interval; GABA = Gamma-aminobutyric Acid; R^2^ = Increment in Nagelkerke’s pseudo-R^2^; *p* = BH adjusted p-value
